# Functional Connectivity Gradients Reveal Altered Hierarchical Cortical Organization in Functional Neurological Disorder

**DOI:** 10.1101/2025.06.05.25328981

**Authors:** Christiana Westlin, Andrew J. Guthrie, Cristina Bleier, Sara A. Finkelstein, Julie Maggio, Jessica Ranford, Julie MacLean, Ellen Godena, Daniel Millstein, Jennifer Freeburn, Caitlin Adams, Christopher D. Stephen, Ibai Diez, David L. Perez, Yuta Katsumi

**Author notes:** Corresponding authors: Christiana Westlin, PhD; Massachusetts General Hospital, 55 Fruit Street, Boston, MA, USA; David L. Perez, MD, MMSc; Massachusetts General Hospital, 55 Fruit Street, Boston, MA, USA. Denotes equally contributing co-senior authors (David L. Perez, Yuta Katsumi).

## Abstract

**Background:** Neuroimaging studies of functional neurological disorder (FND), a condition at the intersection of psychiatry and neurology, often rely on discrete connections or parcellations that may obscure the brain’s functional network architecture. This study applied a gradient-based approach to examine macroscale cortical organization in FND.

**Methods:** We analyzed resting-state functional magnetic resonance imaging (fMRI) data from 64 patients with mixed FND (FND-mixed), 61 age- and sex-matched healthy controls (HCs), and 62 psychiatric controls (PCs) matched on age, sex, depression, anxiety, and post-traumatic stress disorder (PTSD) severity. Functional connectivity gradients were computed to capture dominant axes of cortical organization. Between-group comparisons were conducted for the top three gradients, and associations with symptom severity were investigated. Subtype-specific patterns in functional motor disorder (n=49) and functional seizure (n=24) were also examined. Analyses controlled for age, sex, and antidepressant use, and were *post-hoc* adjusted for depression, anxiety, and PTSD-severity, and for childhood maltreatment.

**Results:** The FND-mixed group showed alterations across all three gradients relative to HCs and PCs. Gradient 1 revealed increased values in sensorimotor regions, reflecting a shift toward more association-like connectivity. Gradient 2 showed altered differentiation between sensory systems. Gradient 3 exhibited reduced functional separation between representational and modulatory regions, with prominent shifts in the anterior cingulate cortex. Several regions displaying between-group differences also showed correlations with FND and somatic symptom severity. Analyses revealed overlapping and distinct patterns across subtypes.

**Conclusions:** We provide novel evidence of atypical hierarchical brain organization in FND, highlighting gradient-based approaches for identifying mechanistically-relevant altered functional brain organization.

## INTRODUCTION

Functional neurological disorder (FND), previously termed conversion disorder, is a condition at the historical origins of modern-day psychiatry and neurology. Patients with FND experience distressing motor, sensory, and cognitive symptoms that are not attributable to macroscopic structural brain lesions (1,2). FND is a prevalent and potentially disabling condition, accompanied by significant personal and societal costs (3–5). Despite having been neglected scientifically throughout the late 20^th^ century compared other neuropsychiatric disorders, earning labels such as “Psychiatry’s Blind Spot” and “Medicine’s Silent Epidemic”, renewed interest in FND has been catalyzed by the use of physical examination signs that enable a highly specific “rule in” diagnostic approach (6–8). Although substantial progress has been made in the past few decades (9,10), the neurobiological mechanisms underlying FND remain incompletely understood (11).

Resting-state functional magnetic resonance imaging (fMRI) studies of FND have revealed alterations across several large-scale brain networks, including the somatomotor, salience, and default mode networks (11,12). These investigations often rely on discrete network parcellations or seed-based analyses, which are useful for summarizing connectivity and testing circuit-level hypotheses (13–22). However, such approaches assume discrete network boundaries or connections, which may obscure the fluid and overlapping nature of functional brain organization. Graph theory methods have also been used to probe alterations in network architecture by modeling the brain as a set of nodes (regions) and edges (connections) (23–27). These approaches have led to insights into the functional brain organization in FND (i.e., increased somatomotor network integrated connectivity (27)), yet they similarly emphasize discrete connections between regions and do not account for more continuous patterns of connectivity.

Gradient mapping offers a complementary perspective for examining macroscale brain organization patterns along multiple continuous dimensions (28–33). These approaches use dimensionality reduction techniques to identify low-dimensional representations of functional connectivity, referred to as *gradients*. When applied to the cerebral cortex, gradients capture smooth transitions in connectivity across the cortex and reflect hierarchical dimensions of cortical organization. Gradient-based analyses have increasingly been used to study macroscale brain alterations in psychiatric and neurological conditions, including depression, autism, schizophrenia, epilepsy, and Alzheimer’s disease (34–38). Research has highlighted three gradients that reflect dominant axes of cortical organization (30,39,40): Gradient 1, typically referred to as the *association-sensorimotor* gradient (also called a ‘transmodal-unimodal’ gradient), is anchored at one end by the default mode network and at the other by sensory and motor networks and the salience network; Gradient 2, typically referred to as the *visual-somatomotor* gradient, distinguishes between visual and somatomotor sensory systems; and Gradient 3, sometimes referred to as the *representation-modulation* gradient, differentiates the default mode and sensory networks from modulatory/attentional (i.e., frontoparietal, dorsal attention, and salience) networks.

These functional connectivity gradients have recently been interpreted through a predictive processing framework of brain function, which hypothesizes that the brain actively constructs predictions about incoming sensory data based on past experiences, rather than passively reacting to sensory input (30,40). Predictive processing offers a compelling model for understanding FND, which has been hypothesized to involve impairments in the construction, weighting, and updating of predictions (41–43). Under this framework, Gradient 1 (association-sensorimotor) is thought to reflect a continuum from regions that are involved in representing abstract, multimodal prediction signals (e.g., default mode network regions) to regions involved in signaling prediction errors (e.g., sensory and motor networks, salience network). Gradient 2 (visual-somatomotor), while requiring more research to aid interpretability, appears to reflect a segregation of exteroceptive sensory systems. Gradient 3 (representation-modulation) can be interpreted as distinguishing regions that represent prediction and prediction error signals on one end (e.g., default mode and sensory networks) from regions that set the precision of these signals through attentional modulation (e.g., salience and frontoparietal networks). As such, observed alterations in these gradients can provide insights into potential disruptions within the predictive hierarchy.

In the present study, we applied a gradient-based framework to investigate alterations in the hierarchical functional organization of the cerebral cortex in patients with FND. We computed connectivity gradients from resting-state fMRI data in a mixed FND cohort (FND-mixed) and compared them to those derived from age- and sex-matched healthy controls (HCs) and psychiatric controls (PCs) matched on age, sex, depression, anxiety, and PTSD symptom severity. Inclusion of a psychiatric control group allowed us to discern whether observed alterations were specific to FND or reflected more general features of co-occurring psychiatric conditions or shared risk factors (44,45). We examined between-group differences in the top three gradients, explored subtype-specific effects in patients with functional motor disorder (FND-motor) and functional seizures (FND-seiz), and evaluated associations between gradient values and both core FND symptom severity and broader somatic symptom burden (46,47). Through these gradient-based analyses, we aimed to advance understanding of large-scale functional brain organization in FND, offering insights into aberrant mechanisms that may inform future biologically-grounded approaches to diagnosis and treatment.

## METHODS

### Participants

Sixty-four participants comprising a mixed cohort of individuals with FND-motor and/or FND-seiz (FND-mixed; 54 female; 10 male; mean age [SD]=40.0±13.8 years; average illness duration=4.2±5.3 years, range=0.3-25 years; **Table 1**) were prospectively recruited from the Massachusetts General Hospital between June 2018 and March 2024 (7,48). FND diagnoses were made based on positive signs, semiological features, and electroencephalography data (functional seizures only (49)). Recruitment included those with FND-motor and/or FND-seiz, given that many individuals present with mixed symptoms, and some people who present with one discrete phenotype commonly develop distinct functional neurological symptoms over the course of their illness (50,51). Participants were excluded if they had major neurological comorbidities (e.g., epilepsy, Parkinson’s disease, etc.), known brain MRI abnormalities, poorly controlled medical problems with central nervous system (CNS) consequences, active illicit substance dependence, known psychosis, and/or active suicidality. The FND-mixed cohort included 49 individuals with FND-motor (tremor=23; weakness=18; gait=17; speech=14; tics/jerks/spasms=10; dystonia=3 [motor phenotypes were not mutually exclusive]) and 24 participants with FND-seiz (documented=17; clinically established=2; probable=5); nine individuals met criteria for both FND-motor and FND-seiz subtypes (**Supplemental Table 1**). Data from this FND cohort have been previously published in one structural MRI study and one graph-theory based fMRI study (27,52).

**Table 1.** Demographic and psychometric characteristics of mixed functional neurological disorder (FND-mixed), psychiatric control (PC) and healthy control (HC) samples.

| | **FND-mixed (N = 64)**<br>Mean ± SD or N | **PCs (N = 62)**<br>Mean ± SD or N (p<br>corrected) | **HCs (N = 61)**<br>Mean ± SD or N (p<br>corrected) |
| --- | --- | --- | --- |
| **Age (years)** | 40.0 ± 13.8 | 35.6 ± 13.2 (0.2) | 36.5 ± 10.9 (0.2) |
| **Sex** | F: 54; M: 10 | F: 52; M: 10 (0.9) | F: 50; M: 11 (0.7) |
| **SDQ-20** | 34.7 ± 15.6 | 22.0 ± 5.2 (<0.001*) | 20.3 ± 0.6 (<0.001*) |
| **PHQ-15** | 13.3 ± 7.3 | 6.3 ± 3.6 (<0.001*) | 2.6 ± 2.2 (<0.001*) |
| **BDI-II** | 16.8 ± 12.3 | 14.6 ± 13.1 (0.4) | 1.5 ± 2.4 (<0.001*) |
| **STAI-Total** | 82.0 ± 29.2 | 79.0 ± 23.6 (0.5) | 53.8 ± 9.2 (<0.001*) |
| **PCL-5** | 28.2 ± 19.6 | 22.0 ± 18.7 (0.1) | 2.6 ± 3.6 (<0.001*) |
| **CTQ-Abuse** | 31.8 ± 14.8 | 26.0 ± 12.0 (0.03*) | 18.0 ± 3.4 (<0.001*) |
| **CTQ-Neglect** | 20.8 ± 9.8 | 19.6 ± 8.8 (0.5) | 13.2 ± 4.2 (<0.001*) |
| **SSRI/SNRI** | 33 | 29 (0.7) | 0 (<0.001*) |

*P* values reflect statistical significance between the FND cohort and the control group after False Discovery Rate (FDR) correction for multiple comparisons. Asterisks indicate corrected *p*-values < 0.05. Statistical comparisons of continuous variables were done using a Mann Whitney U test, while comparisons of discrete variables were done using a Chi-Squared test. Six FND and 1 PC participants had incomplete data. For each variable, counts of missing data were as follows: SDQ-20: 6 FND, 1 PC; PHQ-15: 6 FND; BDI-II: 4 FND; STAI-Total: 4 FND; PCL-5: 5 FND; CTQ: 4 FND. F, Female, M, Male; SDQ-20, Somatoform Dissociation Questionnaire-20; PHQ-15, Patient Health Questionnaire-15; BDI-II, Beck Depression Inventory-II; STAI-Total, Spielberger State-Trait Anxiety Inventory-Total; PCL-5, Post-Traumatic Stress Disorder Checklist for DSM-5; CTQ, Childhood Trauma Questionnaire; SSRI/SNRI, selective serotonin reuptake inhibitor/serotonin norepinephrine reuptake inhibitor use.

Sixty-two PCs (52 female; 10 male; mean age [SD]=35.6±13.2 years) and sixty-one HCs (50 female; 11 male; mean age [SD]=36.5±10.9 years) were prospectively recruited from the community. PCs had a lifetime history of clinically salient depression (*n*=53), anxiety (*n*=43), and/or post-traumatic stress disorder (PTSD; *n=*21) (Forty-two had more than one of these diagnoses; **Supplemental Table 2**). Exclusion criteria for the control cohorts were the same as for FND, with the added exclusion of an FND or somatic symptom disorder diagnosis. HCs had an added exclusion of no history of psychiatric disorders, and no HCs were on psychotropic medications.

Seventeen additional participants (4 FND, 6 PCs, 7 HCs) were enrolled but excluded following imaging acquisition and processing due to excessive head motion (<120 usable volumes). All individuals signed informed consent and the Mass General Brigham Institutional Review Board approved this study.

### Neuropsychiatric Characterization

All participants underwent a Structured Clinical Interview for Diagnostic and Statistical Manual Disorders (SCID-I). Participants also completed a battery of psychometric questionnaires, including the Beck Depression Inventory-II (BDI-II), Spielberger State-Trait Anxiety Inventory (STAI), PTSD Checklist-5 (PCL-5), and Childhood Trauma Questionnaire (CTQ). FND symptom severity was assessed with the Somatoform Dissociation Questionnaire-20 (SDQ-20), a 20-item measure of the extent to which core FND symptoms (e.g., paralysis) were experienced over the past year on a five-point Likert scale (53). Somatic symptom severity was assessed with the Patient Health Questionnaire-15 (PHQ-15), a 15-item measure of how bothersome physical symptoms (e.g., pain, fatigue) were over the past four weeks on a three-point Likert scale (54). Seven participants had incomplete psychometric data (6 FND, 1 PC).

### MRI Acquisition and Preprocessing

3T MRI acquisition details are described in **Supplementary Materials.** Data were preprocessed using FMRIB Software Library v5.0.7 and MATLAB 2023a using in-house preprocessing pipelines (26), as described in **Supplementary Materials.**

### Gradient Analysis

#### Diffusion Map Embedding

Functional connectivity gradients were computed using diffusion map embedding, a nonlinear dimensionality reduction technique that can capture the dominant dimensions of spatial variation in high-dimensional functional connectivity data through a set of low-dimensional manifolds (i.e., gradients) (55,56). To do this, resting state functional connectivity (rsFC) matrices were computed for each participant by calculating Pearson correlation coefficients between the time series of every pair of cortical grey matter voxels. Each participant’s rsFC matrix was thresholded to retain only the top 10% of connections, with all remaining connections set to zero. We then computed a non-negative, square symmetric affinity matrix for each rsFC matrix using cosine similarity to quantify the similarity between connectivity profiles for each pair of voxels. These affinity matrices were used as inputs to diffusion map embedding, which yielded ten gradients per participant, reflecting the dominant dimensions of rsFC spatial variation. Based on *a priori* hypotheses and following prior work, we focused on the three most dominant gradients (30,32,39,40).

To enable statistical comparisons across participants, individual gradients were aligned to group-level template gradients derived from HCs. The template gradients were generated from a group-average rsFC matrix, computed by standardizing each HC’s rsFC matrix via Fisher’s *r*-to-*z* transformation, averaging across participants, and converting the resulting matrix back to Pearson’s *r* values. Group-averaged HC gradients were then computed using the same steps as described above. Individual gradients were then aligned to the HC gradient template using Procrustes rotation (number of iterations=10).

#### Statistical Analysis

General linear models (GLMs) were used to compute between-group differences in voxel-wise gradient values for the top three gradients across FND and HC/PC cohorts. Comparisons were conducted for FND-mixed vs. PCs and FND-mixed vs. HCs separately, followed by subtype analyses for FND-motor and FND-seiz vs. PCs/HCs. All GLMs controlled for age, sex, selective serotonin/norepinephrine reuptake inhibitor (SSRI/SNRI) use (yes/no) and mean framewise displacement (FD). Post-hoc analyses also controlled for: (1) BDI-II, STAI-total, and PCL-5 scores; (2) CTQ-abuse and CTQ-neglect scores. Intersection maps were computed to examine voxels that held across all corrections. To account for multiple comparisons, cluster-wise correction was applied using a Monte Carlo simulation with 10,000 iterations to estimate the probability of false positive clusters at *p*<0.05.

### Symptom Correlations

Post-hoc Spearman correlations were computed between mean gradient values extracted from each significant suprathreshold cluster in FND-mixed vs. PCs comparisons and SDQ-20 and PHQ-15 scores. Correlations with the SDQ-20 were computed for FND-mixed only, while correlations with the PHQ-15 were computed both within FND-mixed only and across FND-mixed and PCs. Multiple comparison correction using false-discovery rate correction was conducted across clusters for each gradient and symptom scale. For significant correlations, post-hoc partial correlation analyses were performed to test if findings held adjusting for FND subtype (i.e., FND-seiz (yes/no)).

## RESULTS

### Demographic and Psychometric Comparisons

There were no differences in age or sex between the FND-mixed, PC, and HC cohorts. Compared to HCs, the FND-mixed group had higher scores on all psychometric scales. Compared to PCs, the FND-mixed group had higher scores on the SDQ-20, PHQ-15, and CTQ-abuse subscale, and did not differ on the BDI-II, STAI-total, PCL-5, CTQ-neglect subscale, or SSRI/SNRI use (see **Table 1** and **Supplementary Table 4**).

### Between-Group Gradient Comparisons

Group-level gradient maps for FND-mixed, HCs, and PCs are shown in **Fig. 1**. Across all participants, the top three gradients collectively accounted for 46.6% of the variance in functional connectivity organization (**Supplementary Fig. 1**). These gradients exhibited high spatial similarity to previously published gradients (30,31; **Supplementary Table 3**), with Gradient 1 (association-sensorimotor) distinguishing default mode and frontoparietal networks from exteroceptive sensory (e.g., somatosensory, visual) and salience networks, Gradient 2 (visual-somatomotor) distinguishing visual from somatosensory/motor networks, and Gradient 3 (representation-modulation) distinguishing default mode and exteroceptive sensory networks from frontoparietal and salience networks.

**Figure 1.**
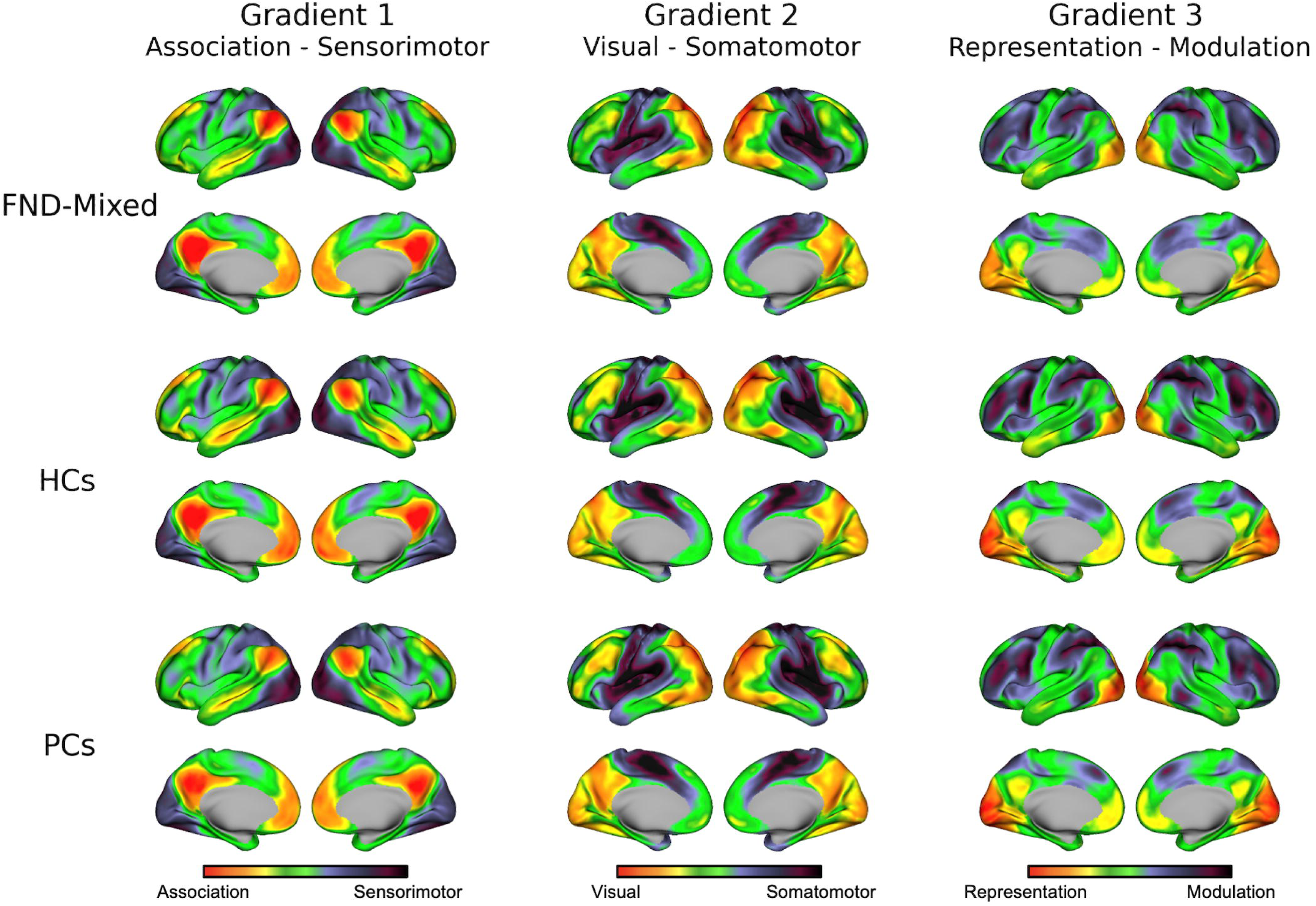
Resting-state functional connectivity gradients of the cerebral cortex for patients with FND (*n*=64; *top*), psychiatric controls (PCs; *n*=62; *middle*), and healthy controls (HCs; *n*=61; *bottom*). Group-averaged patterns are shown for the top three gradients explaining dominant axes along which whole-cortex connectivity is organized (*left,* Gradient 1; *middle,* Gradient 2; *right,* Gradient 3). Voxels with similar connectivity patterns are represented by similar colors along the gradient, while voxels at opposite extremes reflect the greatest dissimilarity in connectivity. The sign of each gradient is arbitrary.

#### Gradient 1 (Association-Sensorimotor)

Compared to HCs, patients in the FND-mixed cohort exhibited increased Gradient 1 values in bilateral precentral and postcentral gyri, slightly extending into the superior and inferior parietal lobules (**Fig. 2, top**). These regions anchor the sensorimotor end of the gradient, such that increases in gradient values for FND-mixed reflect a shift in similarity of functional connectivity profiles towards the association end. These findings held for the primary adjustment of age, sex, SSRI/SNRI use, and motion, as well as for an additional post-hoc correction for BDI-II, STAI-total, and PCL-5 scores (**Supplementary Fig. 2**). However, when correcting post-hoc for CTQ-abuse and CTQ-neglect subscales, only left hemisphere findings remained significant. The FND-mixed group also showed decreased Gradient 1 values relative to HCs in bilateral subgenual anterior cingulate cortex and ventromedial prefrontal cortex, and the right rostral middle temporal gyrus — regions that anchor the association end of the gradient. In this context, decreased values in FND-mixed indicate a shift in similarity of functional connectivity profiles towards the sensorimotor end of the gradient. These decreases, however, did not remain significant following post-hoc corrections.

**Figure 2.**
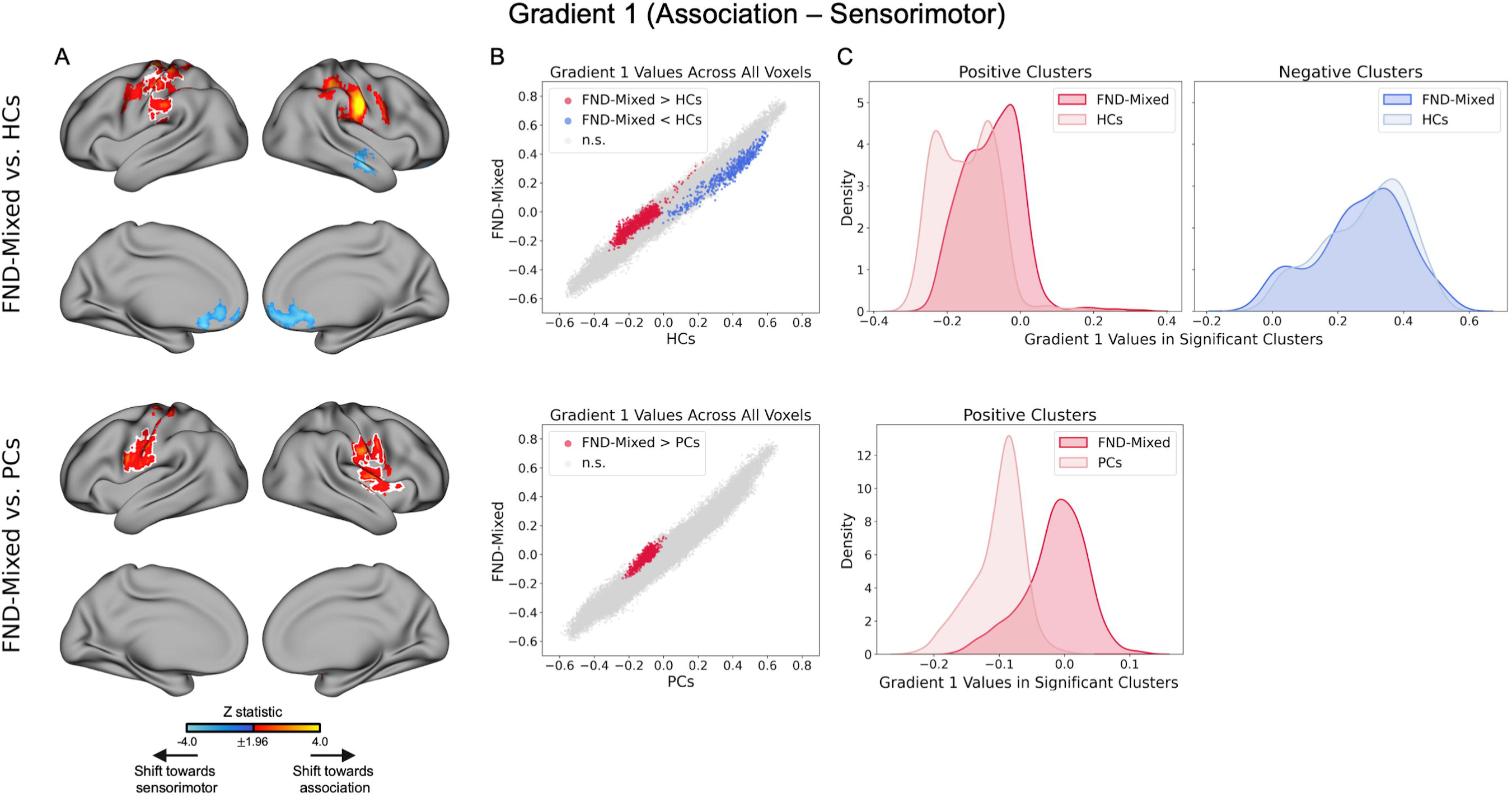
Comparisons of Gradient 1 (association–sensorimotor) for patients with FND-mixed vs. healthy controls (HCs) (*top*) and FND-mixed vs. psychiatric controls (PCs) (*bottom*). (**A**) Results from between-group statistical comparisons. Colors reflect the z-statistic computed from a two-sample general linear model for the primary adjustment for age, sex, SSRI/SNRI use, and mean framewise displacement (z-statistic>1.96; p<0.05 cluster-corrected for multiple comparisons). White outlines reflect regions that also held across all *post-hoc* corrections for: i) BDI-II, STAI-total and PCL-5 scores; and ii) CTQ-abuse and CTQ-neglect scores. Statistical maps for post-hoc comparisons are visualized in Supplementary Figures 2 and 3. (**B**) Scatter plots of gradient values. Each dot represents a cortical voxel. Statistically significant voxels are colored (increases/decreases shown in red/blue, respectively), while non-significant (n.s.) voxels are in gray. (**C**) Density plots of gradient values extracted from statistically significant positive and negative clusters in the primary analysis for FND-mixed (dark red/blue) and respective control groups (light red/blue). BDI-II, Beck Depression Inventory-II; STAI-total, State Trait Anxiety Inventory-Total; PCL-5, PTSD Checklist-5; CTQ, Childhood Trauma Questionnaire.

Compared to PCs, FND-mixed exhibited increased Gradient 1 values in bilateral pre- and postcentral gyri, as well as the right dorsal mid-insula, reflecting a shift from sensorimotor towards the association end of the gradient (**Fig. 2, bottom**). These findings held when correcting post-hoc for (i) BDI-II, STAI-total, and PCL-5 scores, and (ii) CTQ-abuse and CTQ-neglect subscale scores (**Supplementary Fig. 3**). No regions exhibited decreased Gradient 1 values.

#### Gradient 2 (Visual-Somatomotor)

Patients with FND-mixed, compared to HCs, exhibited increased Gradient 2 values in the right middle temporal gyrus and lateral occipital cortex (**Fig. 3, top**). These differences remained significant across all post-hoc corrections (**Supplementary Fig. 2**). These observed increases suggest a shift in similarity of functional connectivity profiles towards, and expansion of, the visual end of the gradient. Conversely, decreases in Gradient 2 values for FND-mixed vs. HCs were observed in the right lateral prefrontal and posterior parietal cortices, reflecting a shift in similarity of functional connectivity profiles from the middle of the gradient towards the somatomotor end. These decreases largely held when adjusting post-hoc for CTQ-abuse and CTQ-neglect but did not hold when adjusting for BDI-II, STAI-total, and PCL-5 scores.

**Figure 3.**
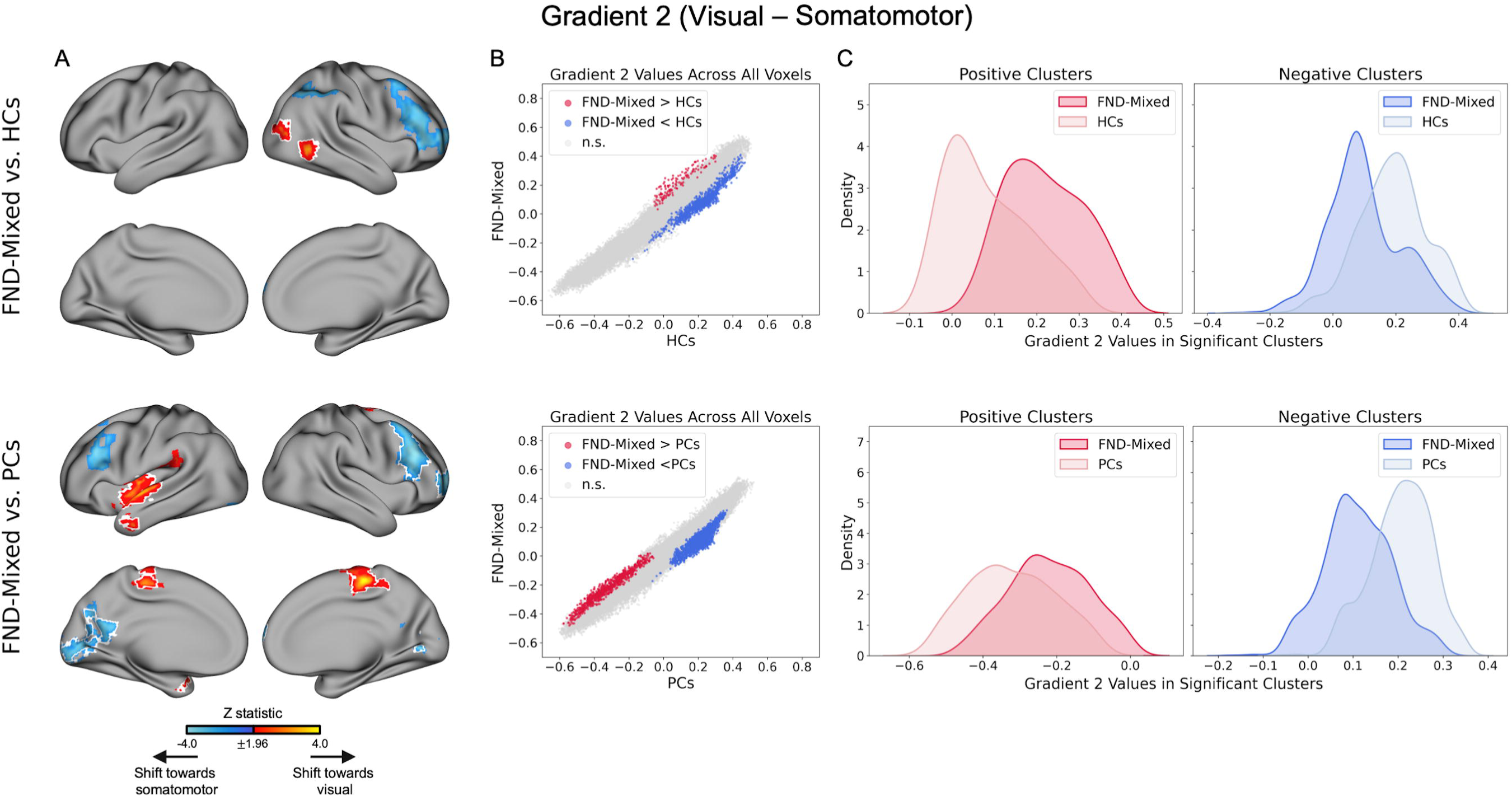
Comparisons of Gradient 2 (visual–somatomotor) for patients with FND-mixed vs. healthy controls (HCs) (*top*) and FND-mixed vs. psychiatric controls (PCs) (*bottom*). (**A**) Results from between-group statistical comparisons. Colors reflect the z-statistic computed from a two-sample general linear model for the primary adjustment for age, sex, SSRI/SNRI use, and mean framewise displacement (z-statistic>1.96; p<0.05 cluster-corrected for multiple comparisons). White outlines reflect regions that also held across all *post-hoc* corrections for: i) BDI-II, STAI-total and PCL-5 scores; and ii) CTQ-abuse and CTQ-neglect scores. Statistical maps for post-hoc comparisons are visualized in Supplementary Figures 2 and 3. (**B**) Scatter plots of gradient values. Each dot represents a cortical voxel. Statistically significant voxels are colored (increases/decreases shown in red/blue, respectively), while non-significant (n.s.) voxels are in gray. (**C**) Density plots of gradient values extracted from statistically significant positive and negative clusters in the primary analysis for FND-mixed (dark red/blue) and respective control groups (light red/blue). BDI-II, Beck Depression Inventory-II; STAI-total, State Trait Anxiety Inventory-Total; PCL-5, PTSD Checklist-5; CTQ, Childhood Trauma Questionnaire.

Compared to PCs, individuals with FND-mixed showed increased Gradient 2 values in bilateral pre-central gyrus/supplementary motor area (SMA), as well as the left middle and superior temporal gyri, temporal pole, mid insula, and parietal operculum (**Fig. 3, bottom**). These regions reflect a shift in similarity of functional connectivity profiles towards the visual end of the gradient, with findings largely remaining significant across post-hoc corrections for (i) BDI-II, STAI-total, and PCL-5 scores and (ii) CTQ-abuse and CTQ-neglect scores (**Supplementary Fig. 3**). Decreases were observed in the bilateral lateral prefrontal and occipital cortices and the left precuneus, with findings remaining significant across post-hoc corrections (decreases in the left prefrontal did not hold when adjusting for CTQ-abuse and neglect). These decreases reflect a shift in similarity of functional connectivity profiles toward the somatomotor end of the gradient.

#### Gradient 3 (Representation-Modulation)

Compared to HCs, patients with FND-mixed exhibited increased Gradient 3 values in bilateral precentral gyri, lateral occipital and superior parietal cortices, and the right post-central, supramarginal, and middle frontal gyri (**Fig. 4, top**). Only the right precentral and middle frontal gyri findings remained significant following both post-hoc corrections (**Supplementary Fig. 2**). These increases reflect a shift in similarity of functional connectivity profiles from the modulation end of the gradient towards the representation end. Decreased Gradient 3 values were observed in the bilateral anterior cingulate, superior frontal, and occipital cortices, as well as the right posterior insula and opercular cortex, reflecting a shift in similarity of functional connectivity profiles from the representation end towards modulation. Decreases in the bilateral anterior cingulate and superior frontal cortices remained significant when correcting for CTQ-abuse and neglect, but no other findings held across post-hoc corrections.

**Figure 4.**
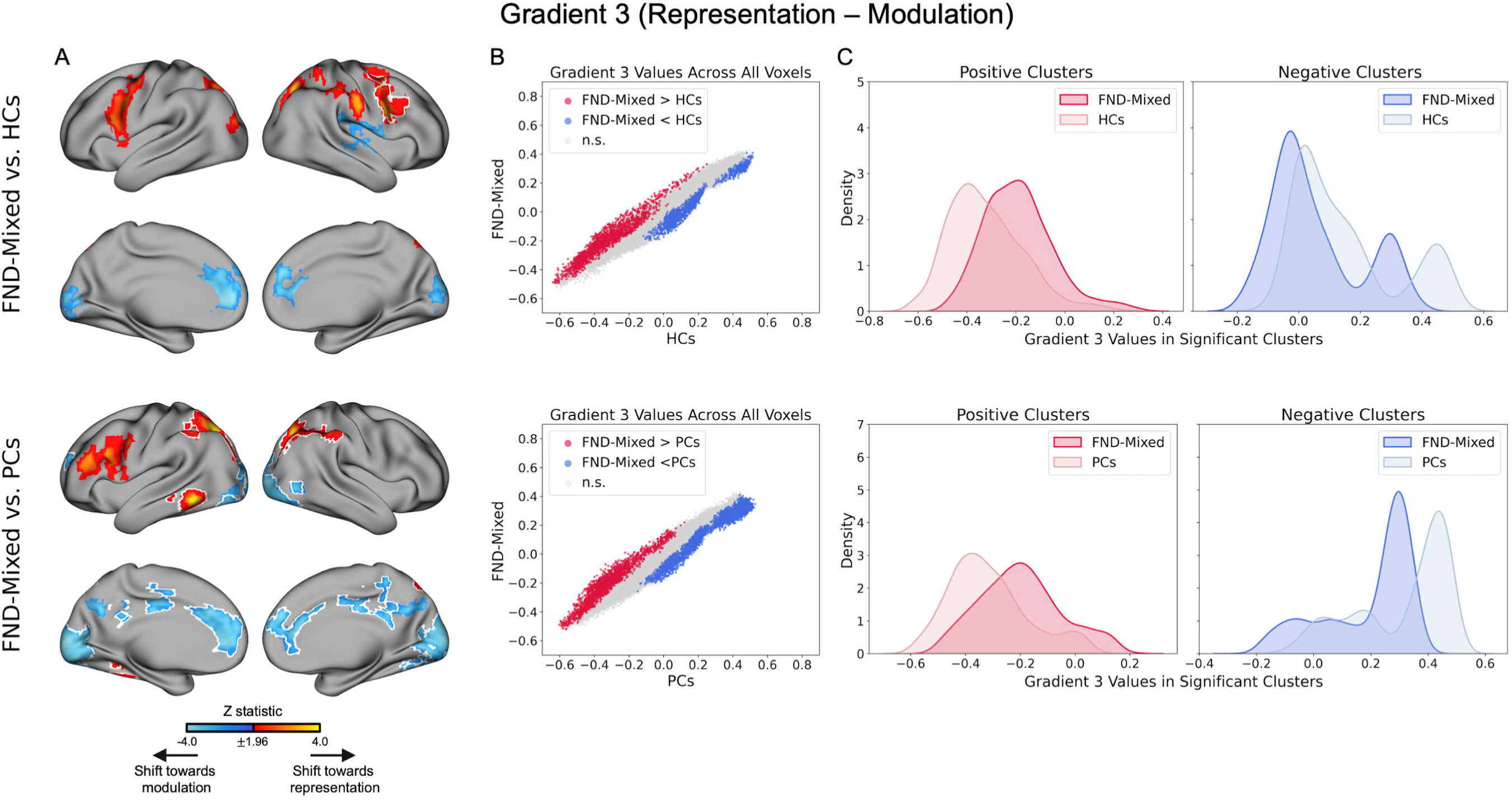
Comparisons of Gradient 3 (representation–modulation) for patients with FND-mixed vs. healthy controls (HCs) (*top*) and FND-mixed vs. psychiatric controls (PCs) (*bottom*). (**A**) Results from between-group statistical comparisons. Colors reflect the z-statistic computed from a two-sample general linear model for the primary adjustment for age, sex, SSRI/SNRI use, and mean framewise displacement (z-statistic>1.96; p<0.05 cluster-corrected for multiple comparisons). White outlines reflect regions that also held across all *post-hoc* corrections for: i) BDI-II, STAI-total and PCL-5 scores; and ii) CTQ-abuse and CTQ-neglect scores. Statistical maps for post-hoc comparisons are visualized in Supplementary Figures 2 and 3. (**B**) Scatter plots of gradient values. Each dot represents a cortical voxel. Statistically significant voxels are colored (increases/decreases shown in red/blue, respectively), while non-significant (n.s.) voxels are in gray. (**C**) Density plots of gradient values extracted from statistically significant positive and negative clusters in the primary analysis for FND-mixed (dark red/blue) and respective control groups (light red/blue). BDI-II, Beck Depression Inventory-II; STAI-total, State Trait Anxiety Inventory-Total; PCL-5, PTSD Checklist-5; CTQ, Childhood Trauma Questionnaire.

Compared to PCs, increased Gradient 3 values for patients with FND-mixed were observed in bilateral lateral occipital and superior parietal cortices and supramarginal gyri, as well as the left middle and inferior frontal gyri, and inferior temporal gyrus (**Fig. 4, bottom**). These increases reflect a shift in similarity of functional connectivity profiles from the modulation end of the gradient towards representation. Most findings remained significant across post-hoc corrections (left frontal findings did not remain significant when correcting post-hoc for CTQ-abuse and neglect; **Supplementary Fig. 3**). Decreased values were observed across bilateral cingulate cortex, as well as bilateral precuneus, superior frontal, and occipital cortices, all of which remained significant across post-hoc corrections. These decreases reflect a shift in similarity of functional connectivity profiles from the representation end of the gradient towards modulation.

### Comparisons Across FND Subtypes

We next examined between-group comparisons for FND-motor and FND-seiz subtypes (vs. HCs and vs. PCs; **Fig. 5**). For Gradient 1, differences observed for the FND-motor cohort were largely the same as the pattern reported for the full FND-mixed cohort. Findings for FND-seiz were less robust, with increases in the post-central gyrus vs. HCs that overlapped with those observed for FND-motor, and no differences observed for FND-seiz vs. PCs. For Gradient 2, a similar pattern emerged where the FND-motor findings were largely the same as the pattern reported for the full FND- mixed cohort. A comparison of FND-seiz vs. HCs revealed no significant differences, and FND-seiz vs. PCs exhibited decreases in the right lateral frontal cortex that overlapped with FND-motor findings. Comparisons for Gradient 3 again revealed a pattern for FND-motor that was largely the same as the pattern reported for the full FND-mixed cohort. Increases in the right post-central gyrus were not observed in FND-motor vs. HCs, but were observed in FND-seiz vs. HCs. Both FND-motor and FND-seiz cohorts revealed overlapping decreases in the bilateral anterior cingulate and superior frontal cortices (vs. HCs and vs. PCs), as well as overlapping increases in the left lateral occipital and superior parietal cortices and decreases in the occipital cortex.

**Figure 5.**
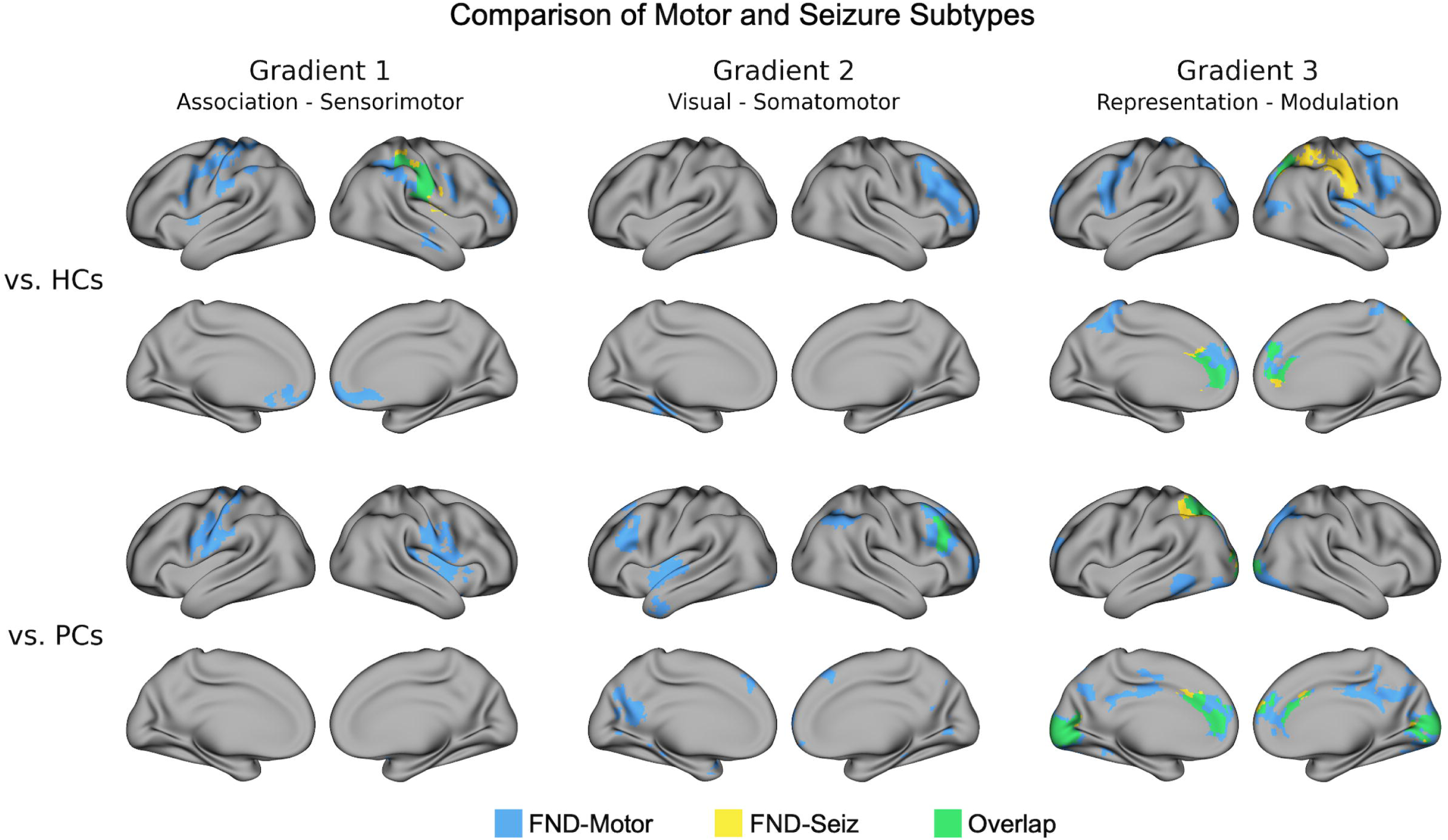
Comparison of functional motor disorder (FND-motor) and functional seizure (FND-seiz) subtypes. Overlays depict voxels that held across all two-sample general-linear models (z-stat>1.96; p<0.05 cluster-corrected for multiple comparisons for Gradient 1 (*left*), Gradient 2 (*middle*), and Gradient 3 (*right*) analyses compared to healthy controls (HCs; *top*) and psychiatric controls (PCs; *bottom*). Results for the FND-motor analyses are shown in blue, FND-seiz analyses shown in yellow, and overlapping regions across both subtype analyses shown in green.

### Symptom Severity Correlations

We observed several significant relationships between self-reported symptoms and gradient values extracted from statistically significant clusters in the FND-mixed vs. PCs analyses. To limit investigations to the most robust findings, only clusters that remained significant for all post-hoc corrections were tested for potential correlations with symptom severity scores. When looking across individuals with FND-mixed and PCs, Gradient 1 values extracted from a cluster in the left pre- and post-central gyri positively correlated with PHQ-15 scores (*r_s_*(115)=0.25, *p_corrected_*=0.01; **Fig. 6a**). Across both FND-mixed and PCs, Gradient 3 values from a cluster comprised of bilateral anterior cingulate and superior frontal cortex negatively correlated with PHQ-15 scores (*r_s_*(114)=-0.28, *p_corrected_*=0.02; **Fig. 6c**). Within FND-mixed alone, Gradient 2 values from a cluster comprised of left mid insula and superior temporal gyri/temporal pole positively correlated with SDQ-20 scores (*r_s_*(54)=0.34, *p_corrected_*=0.04; **Fig. 6b**). All three correlations remained statistically significant in *post-hoc* partial correlation analyses that additionally controlled for FND subtype (FND-seiz, yes/no).

**Figure 6.**
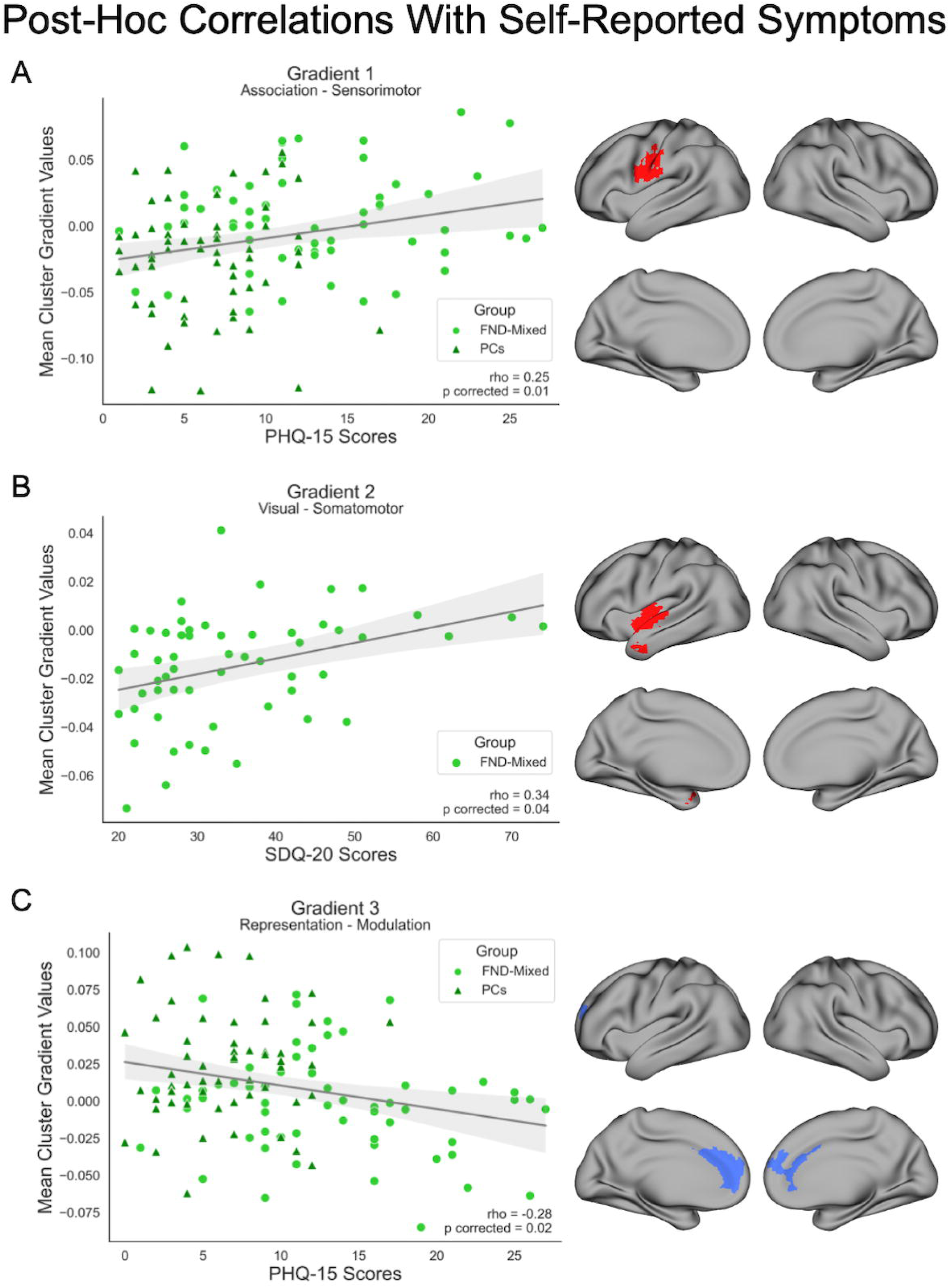
*Post-hoc* correlations between self-reported symptoms and mean gradient values extracted from significant clusters in FND-mixed vs. psychiatric control (PCs) between-group comparisons. (**A**) The significant positive Spearman correlation across FND-mixed and PCs between mean Gradient 1 values extracted from the cluster shown in red and scores on the Patient Health Questionnaire-15 (PHQ-15). (**B**) The significant positive Spearman correlation within FND-mixed participants between mean Gradient 2 values extracted from the cluster shown in red and scores on the Somatoform Dissociation Questionnaire-20 (SDQ-20). (**C**) The significant negative Spearman correlation across FND-mixed and PCs between mean Gradient 3 values extracted from the cluster shown in blue and scores on the PHQ-15. Shaded regions reflect 95% confidence intervals. Outliers were removed prior to computing the correlation coefficients if they had a mean value that exceeded more than 1.5 times the interquartile range above the upper quartile or below the lower quartile. Colored regions on brain maps indicate clusters with significant increases (red) or decreases (blue) in FND-mixed vs. PCs comparisons. For scatterplots, FND-mixed participants are depicted with light green circles and PCs with dark green triangles.

## DISCUSSION

Using a gradient-based approach to investigate macroscale functional brain organization, we observed significant alterations across all three gradients in the FND- mixed group relative to controls, several of which were associated with individual differences in symptom severity. Notably, Gradient 1 showed connectivity shifts in sensorimotor regions towards more association-like profiles, Gradient 2 revealed altered visual-somatomotor differentiation, and Gradient 3 exhibited reduced separation in rsFC profiles between representational and modulatory regions. We also observed both overlapping and distinct alterations in FND-motor and FND-seiz subtypes. Together, these findings provide convergent evidence for disrupted hierarchical functional brain organization in FND. Although our findings are derived from fMRI-based rsFC and thus are inherently correlational, we interpret our findings through the lens of predictive processing accounts of brain function, potentially reflecting aberrant predictive mechanisms in FND (57,58).

Gradient 1, the association-somatomotor gradient, spans from higher-order association regions (e.g., default mode network) to ‘unimodal’ sensorimotor regions, and has been proposed to reflect a continuum between regions involved in representing prediction signals to those involved in signaling prediction errors (30,40). In the FND- mixed cohort, increased Gradient 1 values were observed in the pre- and postcentral gyri, indicating that these sensorimotor regions exhibited more association-like connectivity profiles. These increases were present compared to both HCs and PCs, and largely remained significant after *post-hoc* adjustments for psychiatric symptoms and trauma burden, underscoring their specificity to FND. From a predictive processing perspective, these findings suggest that regions typically involved in signaling prediction errors are instead exhibiting connectivity profiles similar to areas involved in representing predictions, which is consistent with theories that have proposed overly strong priors in FND that are not properly updated based on incoming sensory inputs (41–43). These findings are also consistent with recent evidence from our group of increased somatomotor integration (i.e., between-network connectivity) in FND-mixed (27), as well as enhanced link-step connectivity from primary motor cortex to the posterior insula and mid cingulate cortex (23). The current gradient-based approach extends prior findings by revealing not just which connections are altered, but *how* their functional roles are reorganized within the broader connectivity landscape, offering a macroscale perspective of disrupted organization in FND. Additionally, reinforcing the clinical relevance of these findings, Gradient 1 values in the left pre- and postcentral gyri were significantly correlated with self-reported somatic symptoms across both patients with FND-mixed and PCs. This is noteworthy given that somatic (non-motor) symptoms are strongly associated with reductions in health-related quality of life in this population (59), and have been reported to precede onset of core FND symptoms by more than six years (60).

Gradient 2, the visual-somatomotor gradient, is thought to reflect the organization of sensory systems. In the FND-mixed cohort, decreased Gradient 2 values were observed in lateral prefrontal cortices relative to both HCs and PCs, suggesting that attentional regions, positioned towards the middle of the gradient, are shifting towards more somatomotor-like connectivity profiles. This may reflect abnormal attentional modulation of sensory systems, such that maladaptive predictions are not properly updated based on incoming sensory inputs. The FND-mixed cohort also exhibited increased Gradient 2 values in somatomotor regions compared to PCs, indicating a shift towards more visual-like connectivity that may reflect aberrant processing of sensory signals. Of note, Gradient 2 values in the left mid-insula were positively correlated with self-reported core functional neurological symptoms within the FND-mixed cohort. These findings align with the growing literature of a fundamental role for altered sensory processing in FND (57,61,62).

Gradient 3, the representation-modulation gradient, differentiates regions involved in representing predictions and prediction errors (e.g., default mode and sensory networks) from those involved in modulating their precision (e.g., salience and frontoparietal networks). In the FND-mixed cohort compared to both HCs and PCs, increased Gradient 3 values were observed in modulatory regions and decreased values in representation regions, suggesting less functional distinction between these processes. This pattern may reflect atypical predictive and precision-setting processes, potentially leading to dominant priors and impaired updating that have been hypothesized in FND (41–43). Notably, the decrease in Gradient 3 values within the anterior cingulate cortex (perigenual and dorsal regions) was robustly found relative to PCs and remained significant after adjusting for depression, anxiety, PTSD symptoms, and trauma burden, indicating specificity to FND beyond shared pathology and risk factors. Moreover, these values were negatively associated with somatic symptom severity across FND-mixed and PCs, underscoring the clinical relevance of altered connectivity profiles in regions involved in precision setting. Among other considerations, these findings also support the need for future research targeting the non-invasive modulation of the perigenual/dorsal anterior cingulate cortex as a potential treatment for disabling somatic symptoms (63).

Analyses of FND subtypes revealed shared and distinct alterations across the top three connectivity gradients. The FND-motor cohort showed patterns that were largely consistent with those observed in the full FND-mixed cohort, while findings for the FND-seiz group were less robust. For Gradients 1 and 2, there was limited overlap between FND-motor and FND-seiz cohorts, with both groups exhibiting Gradient 1 increases in the right postcentral gyrus (vs. HCs) and Gradient 2 decreases in the right lateral frontal gyrus (vs. PCs). Gradient 3 exhibited the most overlap between subtypes, with convergent decreases in anterior cingulate and occipital cortices (vs. HCs and PCs). These results highlight the neurobiological heterogeneity across FND subtypes, alongside shared disruptions in cortical organization.

Some alterations were also observed only in comparisons between the FND- mixed group and HCs but not present for comparisons against PCs. These findings, while still indicative of disrupted functional organization, may reflect features that are not specific to FND but instead are shared across psychiatric conditions. For example, decreased Gradient 1 values in FND-mixed vs. HCs in the ventromedial prefrontal and subgenual anterior cingulate cortices did not survive *post-hoc* corrections, and were not present in comparisons against PCs, suggesting that while association-level disruptions may occur, their relation to FND-specific pathophysiology is less clear. Similarly, we observed increases in visual regions for Gradient 2, reflecting a potential expansion of the gradient, that was only present in comparisons against HCs. The increases observed in Gradient 3, while spanning modulatory regions across both control group comparisons, were also unique for HC comparisons compared to those with PCs. These findings may reflect more general vulnerabilities or comorbidities, and thus their role in FND’s specific pathophysiology remains less clear.

This study has several limitations. While we interpret many of the observed findings as likely disorder-related, due to their persistence after adjusting for covariates and their presence in comparisons with both HCs and PCs, they may also reflect compensatory processes rather than core pathophysiology. Additionally, although the PCs were matched on severity of key mental health dimensions, diagnostic heterogeneity or unmeasured clinical variables may still have influenced the findings. While between-group findings were contextualized based on their additional associations with self-reported symptom severity measures (a study strength), future research should also aim to incorporate the use of clinician-rated and objective measures of severity (64). The smaller sample sizes in subtype cohorts, particularly FND-seiz, may also have limited the ability to detect more subtle effects. Future research should also seek to reconcile how the shifts in rsFC profiles observed in this study relate to subtle structural alterations in grey and white matter previously observed in FND cohorts (52,65,66). Moreover, our analysis focused exclusively on cortical gradients due to methodological limitations of including subcortical structures, including the small number of voxels and correspondingly lower variance these regions contribute to whole-brain connectivity patterns; future work should explore functional gradients within the subcortex to provide a more complete picture of macroscale organization in FND. Finally, although we have interpreted the findings within a predictive processing framework, the overall functional implications of gradient-based alterations warrant further exploration. The absence of symptom correlations in several regions showing group-level differences does not preclude functional relevance, but may instead reflect the complex, nonlinear nature of system-level disruptions in FND, highlighting the need for future investigations grounded in a complex systems framework (67).

In conclusion, this study provides compelling evidence that FND is associated with alterations in macroscale cortical organization. We identified shifts across the top three functional connectivity gradients, reflecting disruptions in the hierarchical brain architecture that support the generation of predictions, the updating of these predictions based on sensory input, and the modulation of attention toward or away from these signals. The inclusion of PCs underscores that these alterations are not solely attributable to concurrent mental health symptoms, supporting their FND specificity. Moreover, associations between gradient values in key regions with self-reported FND symptoms and more general somatic symptom severity highlight the pathophysiological relevance of altered functional organization. Future work should aim to replicate and extend these findings in larger, clinically diverse samples, with the goal of informing biologically grounded approaches to diagnosis and treatment.

## DATA AVAILABILITY

For qualified researchers, analysis code and de-identified data pertaining to study results can be made available following local IRB approval upon reasonable request.

## ACKNOWLEDGMENTS

We thank all the research participants, including those with FND, for their participation.

