## Supplementary Materials for "Functional Connectivity Gradients Reveal Altered Hierarchical Cortical Organization in Functional Neurological Disorder"

**MRI Acquisition and Preprocessing**

Participants were scanned on the same Siemens Tim Trio 3T MRI scanner using a 12-channel phased-array head coil. A high-resolution T1-weighted magnetization-prepared rapid gradient echo (MP-RAGE) scan was acquired for each subject with the following parameters: 1mm isotropic voxels; 160 sagittal slices; acquisition matrix size=256x256; repetition time=2300ms; echo time=2.98ms; field of view=256mm. Resting-state blood-oxygen-level-dependent functional scans were acquired using T2*-weighted echo-planar imaging sequences with the following parameters: TR=3000ms; TE=30ms; flip angle 85; 216mm FOV; 3mm isotropic voxels; sequence length=6 minutes, 12 seconds (124 time points/scan). All participants with FND, and all but 26 control participants, had two functional acquisitions (5 PCs, 21 HCs had one scan). Participants were instructed to remain as still as possible with their eyes open, bi-temporal foam pads restricted head motion, and earplugs were used to attenuate scanner noise.

Preprocessing for anatomical and functional MRI data was done using FMRIB Software Library v5.0.7 (FSL, Oxford, UK) and MATLAB 2023a (MathWorks, Natick, MA) with in-house preprocessing pipelines that have been described previously (26,27). Preprocessing of T1-weighted anatomical images included: reorientation to right-posterior-inferior (RPI); alignment to the anterior and posterior commissures; skull stripping; segmentation of grey matter, white matter, and cerebrospinal fluid; and computation of non-linear transformation between individual skull-stripped T1 images and a 3mm resolution MNI152 template. fMRI preprocessing steps included: discarding the first four volumes; slice timing correction; reorientation to RPI; realignment of functional volumes via a 6-parameter rigid body transformation; computation of the transformation between individual skull-stripped T1 images and mean functional images; intensity normalization; and regression of nuisance signals, including 12 motion-related covariates (rigid motion parameters and their derivatives), linear and quadratic terms, and five components each from the lateral ventricles and white matter. Further steps included transformation to MNI space, spatial smoothing using a 3mm full-width at half-maximum Gaussian kernel, and temporal band-pass filtering (0.01-0.08Hz). Head motion was quantified using realignment parameters, including three translation and three rotation estimates, and volumes with excessive head motion (framewise displacement >0.5mm) were removed from the data. Analyses were performed on 120 time points per subject; for those with more than one resting-state acquisition, the runs were concatenated, and the first 120 time points were retained to standardize the time series length across participants.

**SUPPLEMENTARY FIGURE CAPTIONS**

**Supplementary Figure 1.** Scree plot illustrating the proportion of variance explained by each of the top ten functional connectivity gradients.

**Supplementary Figure 2.** Results from between-group statistical comparisons of FND-mixed vs. healthy controls (HCs) across the three gradients (*left,* Gradient 1; *middle,* Gradient 2; *right,* Gradient 3). The top row depicts results from the primary adjustment for age, sex, SSRI/SNRI use, and head motion (mean framewise displacement), the middle row depicts results from the *post-hoc* adjustment controlling additionally for BDI-II, STAI-total, and PCL-5 scores, and the bottom row depicts results from the *post-hoc* adjustment controlling additionally for CTQ-abuse and CTQ-neglect scores. Colors reflect the z-statistic computed from a two-sample general linear model (z-statistic>1.96; p<0.05 cluster-corrected for multiple comparisons). BDI-II, Beck Depression Inventory-II; STAI-total, State Trait Anxiety Inventory-Total; PCL-5, PTSD Checklist-5; CTQ, Childhood Trauma Questionnaire.

**Supplementary Figure 3.** Results from between-group statistical comparisons of FND-mixed vs. psychiatric controls (PCs) across the three gradients (*left,* Gradient 1; *middle,* Gradient 2; *right,* Gradient 3). The top row depicts results from the primary adjustment for age, sex, SSRI/SNRI use, and head motion (mean framewise displacement), the middle row depicts results from the *post-hoc* adjustment controlling additionally for BDI-II, STAI-total, and PCL-5 scores, and the bottom row depicts results from the *post-hoc* adjustment controlling additionally for CTQ-abuse and CTQ-neglect scores. Colors reflect the z-statistic computed from a two-sample general linear model (z-statistic>1.96; p<0.05 cluster-corrected for multiple comparisons). BDI-II, Beck Depression Inventory-II; STAI-total, State Trait Anxiety Inventory-Total; PCL-5, PTSD Checklist-5; CTQ, Childhood Trauma Questionnaire.

**Supplementary Table 1. Demographic characteristics of participants with functional neurological disorder.**

| **FND Subject** | **FND**  **Subtype** | **Phenotypic Description** | **Current SCID-I**  **Diagnoses** | **Past SCID-I Diagnoses** | **Psychotropic Medications** |
| --- | --- | --- | --- | --- | --- |
| 1 | FND-Seiz | documented functional seizures | - | MDE, PTSD | DLX, LTG |
| 2 | FND-Motor | clinically-established functional tics | ANX NOS | - | CTP |
| 3 | FND-Motor | clinically-established functional tremor, functional gait, & functional limb weakness (left hand, right foot) | PTSD, GAD, Eating Disorder | BPAD-II (with MDE) | BUP, BSP, CLP, GBP, LTG |
| 4 | FND-Motor | clinically-established functional tremor, functional jerks, & functional speech | ANX NOS, Somatoform Pain Disorder | PTSD | GBP, LTA |
| 5 | FND-Motor | clinically-established tremor | Specific Phobia, ANX NOS | - | - |
| 6 | FND-Motor | clinically-established functional limb weakness (legs) | DYS, Somatization Disorder, GAD | MDE | DLX, BUP, SERT |
| 7 | FND-Seiz, FND-Motor | documented functional seizures, clinically-established functional tremor & functional speech | AG, Social Phobia, Somatoform Pain Disorder, Undifferentiated Somatoform Disorder | MDE | CLP, AMT |
| 8 | FND-Seiz, FND-Motor* | clinically-established functional seizures & clinically-established functional tremor | - | PTSD | - |
| 9 | FND-Seiz | documented functional seizures | - | MDE | SERT |
| 10 | FND-Motor* | clinically-established functional gait | ANX NOS, Undifferentiated Somatoform Disorder | DEP NOS, Specific Phobia | - |
| 11 | FND-Motor* | clinically-established functional limb weakness (left arm/leg) | DYS, MDE, GAD, PD+AG, Somatization Disorder, Hypochondriasis | PTSD | - |
| 12 | FND-Seiz, FND-Motor | documented functional seizures; clinically-established functional limb weakness (left leg) | - | MDE, PTSD, PD-AG | LTG, TZD |
| 13 | FND-Motor | clinically-established functional tremor | Somatoform Pain Disorder | ETOH Abuse, ANX NOS | DLX, CLP |
| 14 | FND-Seiz | clinically-established functional seizures | MDE, PD-AG, PTSD | MDE | ECP, QTP |
| 15 | FND-Motor | clinically-established functional limb weakness (arms/legs) | ANX NOS | MDE, PTSD, Eating Disorder | TZD, ECP, BUP |
| 16 | FND-Motor | clinically-established functional jerks, & functional limb weakness (legs) | MDE, PD+AG, PTSD, GAD, Somatoform Pain Disorder | MDE | ECP |
| 17 | FND-Motor, FND-Seiz | clinically-established functional tremor, functional gait, functional jerks/spasms, probable functional seizures | PD+AG, DEP NOS | - | BTP, CLP, DVX, HDZ |
| 18 | FND-Motor | clinically-established functional tremor & functional gait | MDE, SAD | MDE, PD+AG | FLX, CLP |
| 19 | FND-Motor | clinically-established functional tremor & functional gait | DYS, GAD, AG | Eating Disorder | SERT |
| 20 | FND-Seiz | documented functional seizures | PTSD | - | HDZ, ECP, GBP, PZN |
| 21 | FND-Seiz | documented functional seizures | GAD | MDE, PTSD | ECP, DZP |
| 22 | FND-Motor | clinically-established functional gait & functional speech | PTSD, Eating Disorder | MDE | QTP, ECP, CLP, HDZ, PZN |
| 23 | FND-Motor* | clinically-established functional limb weakness (left arm/leg) & functional gait | DYS, PD+AG, PTSD, SSD | MDE, Eating Disorder | LDA, CLP, ECP, QTP, BCP |
| 24 | FND-Seiz | documented functional seizures | DEP NOS | MDE, AG | NRT, LTG |
| 25 | FND-Motor* | clinically-established functional tremor | GAD, IAD | MDE, PTSD | DLX, LRZ |
| 26 | FND-Motor | clinically-established functional jerky movements | GAD | Eating Disorder, AUD, SUD | ECP, PGB, AMT, APM / DXAM |
| 27 | FND-Motor | functional tremor | ANX NOS | - | GBP |
| 28 | FND-Motor | clinically-established functional jerky movements | GAD | DEP NOS | SERT |
| 29 | FND-Seiz | documented functional seizures | - | DEP NOS, SAD, ANX NOS | LRZ |
| 30 | FND-Seiz | documented functional seizures | PTSD, GAD | MDE | BSP |
| 31 | FND-Motor | clinically-established functional limb weakness (left arm) & functional speech | GAD, SSD | MDE, PTSD | CBD |
| 32 | FND-Seiz | probable functional seizures | ANX NOS | PTSD, GAD, MDE | SERT, TZD, PZN |
| 33 | FND-Seiz | probable functional seizures | - | ANX NOS | - |
| 34 | FND-Seiz, FND-Motor | clinically-established functional tremor, probable functional seizures | - | MDE | AMT |
| 35 | FND-Seiz, FND-Motor | documented functional seizures, clinically-established functional limb weakness (left leg) & functional gait | MDE, GAD, PD+AG | MDE | CLP, FLX, PZN |
| 36 | FND-Seiz | documented functional seizures | GAD | DEP NOS, AUD | FLX, APZ |
| 37 | FND-Motor* | clinically-established functional tremor & functional speech | PTSD, GAD, PD+AG, MDE | - | DLX, BUP, LRZ |
| 38 | FND-Motor | clinically-established functional gait & functional speech | AG, ADHD | - | APZ, CBD |
| 39 | FND-Seiz, FND-Motor | documented functional seizures; clinically-established functional gait & functional speech | ANX NOS | PTSD, PD+AG | PGB, CBD |
| 40 | FND-Motor | clinically-established functional gait, functional tremor, functional limb weakness (right arm/leg), & functional speech | GAD, PD+AG | PTSD, MDE | TPM |
| 41 | FND-Motor* | clinically-established functional gait | ANX NOS, SSD | GAD, MDE | SERT, LRZ, GBP |
| 42 | FND-Motor* | clinically-established functional gait & functional limb weakness (right arm/leg) | - | MDE, PTSD, Eating Disorder | GBP, AMT, TZD, PZN |
| 43 | FND-Motor | clinically-established functional tremor | - | PTSD, BPAD-II (with MDE) | LTM, QTP, LRZ |
| 44 | FND-Motor | clinically-established functional tremor, functional dystonia & functional gait | PD+AG, SSD | GAD, DEP NOS | - |
| 45 | FND-Motor | clinically-established functional limb weakness (legs) & functional speech | - | DEP NOS, Specific Phobia | - |
| 46 | FND-Motor | clinically-established functional facial spasms/tics & functional speech | GAD, PD+AG, ADHD, PTSD | MDE | CLN, LDA, APR, SERT, LRZ |
| 47 | FND-Motor | clinically-established functional gait & functional speech | - | DEP NOS, ANX NOS | GBP |
| 48 | FND-Motor | clinically-established functional limb weakness (left leg) | ADHD | DYS, MDE, GAD, SAD | DLX, LDA, PGB |
| 49 | FND-Motor | clinically-established functional jerks/spasms/tics | PTSD, ADHD | SAD, MDE, Specific Phobia | MIR, SERT, MPD, LRZ |
| 50 | FND-Seiz | documented functional seizures | GAD, ADHD | MDE, AG | LTG, NRT, FLX, MPD |
| 51 | FND-Motor | clinically-established functional limb weakness (right arm/leg) & functional speech | PTSD, GAD, MDE, PD+AG, SSD | MDE | APR, FLX, GBP, LRZ, MLT |
| 52 | FND-Motor | clinically-established functional tremor & functional dystonia (right foot) | MDE, GAD, PTSD | MDE, PD+AG, SAD | BSP |
| 53 | FND-Seiz | documented functional seizures | GAD, PTSD, ADHD | Eating Disorder, MDE, PD+AG | OLZ, CTP, APM / DXAM |
| 54 | FND-Seiz | documented functional seizures | MDE, GAD | MDE, SAD, Eating Disorder | LTG, LRZ |
| 55 | FND-Motor | clinically-established functional limb weakness (left leg) | ANX NOS | MDE, ANX NOS | GBP, HDZ, ECP |
| 56 | FND-Seiz, FND-Motor | probable functional seizures; clinically-established functional limb weakness (bilateral leg), functional tremor, & functional gait | SSD, AG, DYS | MDE, PD-AG, GAD, PTSD, Eating Disorder | AMT, DLX |
| 57 | FND-Motor | clinically-established functional limb weakness (4-limb) & functional tremor | SSD, SAD, GAD, PD | PTSD, ADHD | SERT, CLP |
| 58 | FND-Motor | clinically-established functional dystonia & functional speech | GAD | PTSD | - |
| 59 | FND-Motor | clinically-established functional limb weakness (left arm & leg), functional tremor, & functional jerks | - | PTSD | - |
| 60 | FND-Motor | clinically-established functional tremor | PTSD, MDE Anxiety NOS | - | MIR, CLP |
| 61 | FND-Motor | clinically-established functional tremor* | PTSD, MDE, SAD, GAD, SSD | DYS, PTSD, AUD, ANX NOS | APZ, BSP; PZN, ECP, TZD, HDZ |
| 62 | FND-Seiz, FND-Motor | documented functional seizures; clinically-established* functional jerks/tics, & functional speech | PD+AG, DYS, GAD, SSD | OCD | DLX, GBP, LRZ |
| 63 | FND-Seiz | documented functional seizures | AG, SAD, OCD, SSD | - | HDZ, DZP |
| 64 | FND-Motor | clinically-established functional tremor, & functional gait | Anxiety NOS | - | HDZ |

*Indicates subject also had concurrent functional somatosensory loss (e.g., non-dermatomal somatosensory deficits). Subjects 1-19 were evaluated using a SCID-I for DSM-IV-TR, while subjects 20-64 were evaluated using the SCID-I for DSM-5; subject 20 had missing SCID-I data and psychiatric comorbidities are based on chart-review diagnoses. FND-Motor, Functional Motor Disorder; FND-Seiz, Functional Seizures; ADHD, Attention-Deficit/Hyperactivity Disorder; AG, Agoraphobia; ANX, Anxiety; AUD, Alcohol Use Disorder; BPAD, Bipolar Affective Disorder; DEP, Depression; DYS, Dysthymia; ETOH-Abuse, Alcohol Abuse; GAD, Generalized Anxiety Disorder; IAD, Illness Anxiety Disorder; MDE, Major Depressive Episode; NOS, not otherwise specified; OCD, Obsessive Compulsive Disorder; PD+AG, Panic Disorder with Agoraphobia; PD-AG, Panic Disorder without Agoraphobia; PTSD, Post-Traumatic Stress Disorder; SAD, Social Anxiety Disorder; SSD, Somatic Symptom Disorder; SUD, Substance Use Disorder; AMT, Amitriptyline; APM, Amphetamine; APR, Aripiprazole; APZ, Alprazolam; BCP, Baclophen; BSP, Buspirone; BTP, Benztropine; BUP, Bupropion; CBD, Cannabidiol; CLN, Clonidine; CLP, Clonazepam; CTP, Citalopram; DLX, Duloxetine; DVX, Desvenlafaxine; DXAM, Dextroamphetamine; DZP, Diazepam; ECP, Escitalopram; FLX, Fluoxetine; GBP, Gabapentin; HDZ, Hydroxyzine; LDA, Lisdexamfetamine; LTM, Lithium; LTG, Lamotrigine; LRZ, Lorazepam; LTA, Levetiracetam; MIR, Mirtazapine; MLT, Melatonin; MPD, Methylphenidate; NRT, Nortriptyline; OLZ, Olanzapine; PGB, Pregabalin; PZN, Prazosin; QTP, Quetiapine; SERT, Sertraline; TPM, Topiramate; TZD, Trazodone.

**Supplementary Table 2. Demographic characteristics of psychiatric controls with a lifetime history of clinically-salient depression, anxiety and/or post-traumatic stress disorder.**

| **PC**  **Subject** | **Current SCID-I**  **Diagnoses** | **Past SCID-I**  **Diagnoses** | **Psychotropic Medications** |
| --- | --- | --- | --- |
| 1 | - | MDE, PTSD | - |
| 2 | - | DEP NOS | - |
| 3 | - | PTSD | - |
| 4 | - | PTSD | - |
| 5 | - | DEP NOS | - |
| 6 | - | ANX NOS | - |
| 7 | MDE | MDE | PAX, BUP |
| 8 | GAD | MDE, PD+AG, Eating Disorder | CTP |
| 9 | DEP NOS | PTSD, MDE | SERT, LTG, TZD, QTP |
| 10 | DYS | MDE | BUP |
| 11 | ANX NOS | MDE | BUP, SERT |
| 12 | GAD, MDE, Eating Disorder | PD-AG, PTSD | BUP, LTG, LRZ, LTM, TZD |
| 13 | MDE | Specific Phobia | - |
| 14 | - | PTSD, DEP NOS | CTP |
| 15 | DYS, ANX NOS | MDE, PD+AG | BUP, SERT |
| 16 | MDE, Eating Disorder, PTSD, GAD | - | DLX |
| 17 | ANX NOS | ANX NOS, DEP NOS, Eating Disorder, ETOH Abuse | VEN |
| 18 | BPAD-II (current mild depression) | MDE | DLX, LTG, QTP |
| 19 | DEP NOS, PD+AG, PTSD, Social Phobia | MDE | DLX, CLP, LTG, QTP |
| 20 | MDE | MDE, ANX NOS | BUP, DLX |
| 21 | MDE, PTSD | MDE, AUD | ZPD, MPD, TZD, MIR, GBP, TPM |
| 22 | GAD, DEP NOS | MDE, PD-AG | ECP |
| 23 | MDE, GAD | PTSD | BUP, LTG, BSP |
| 24 | GAD | MDE, Eating Disorder, Anxiety NOS | FLX, LRZ |
| 25 | MDE, GAD | PTSD | FLX, MIR |
| 26 | MDE, DYS, GAD, AG | - | ECP, APR |
| 27 | - | MDE, ANX NOS | - |
| 28 | - | ANX NOS, DEP NOS | VEN, BUP |
| 29 | ANX NOS | MDE | - |
| 30 | DEP NOS, ANX NOS | MDE | SERT, QTP, GBP |
| 31 | GAD | MDE | BUP, MLT |
| 32 | DYS, ANX NOS | MDE, AUD | BSP, SERT |
| 33 | GAD, SAD | - | SERT |
| 34 | MDE, PTSD, GAD, OCD | - | BSP, LTG, BUP, APM / DXAM, TZD, MLT |
| 35 | Eating Disorder | MDE, PD-AG, OCD | - |
| 36 | MDE, GAD, Specific Anxiety | MDE, PTSD, OCD, Eating Disorder | SERT, LTG, ECP, TZD, MLT |
| 37 | PTSD, SAD, Eating Disorder, ADHD | MDE | TPM, APM / DXAM |
| 38 | MDE, ANX NOS | MDE | LTG, TZD, LRZ |
| 39 | ANX NOS | DEP NOS, GAD | SERT |
| 40 | GAD | - | ECP, BUP, APZ |
| 41 | - | GAD, DEP NOS | BUP |
| 42 | - | ANX NOS | SERT |
| 43 | - | MDE | - |
| 44 | - | MDE, Specific Phobia | - |
| 45 | DEP NOS, ANX NOS | MDE | DLX, BUP |
| 46 | - | MDE, PD+AG | - |
| 47 | - | DEP NOS | - |
| 48 | - | PTSD, MDE | - |
| 49 | GAD | MDE | - |
| 50 | - | DEP NOS | - |
| 51 | - | MDE, PTSD, PD+AG, OCD, AUD | - |
| 52 | PTSD, SAD, ADHD, PD+AG | MDE, Eating Disorder | LDA |
| 53 | - | PTSD | - |
| 54 | - | MDE | GBP, CTP |
| 55 | GAD, MDE, SAD, Specific Phobia | DYS, PTSD | CLP |
| 56 | PTSD | MDE | ECP, CLN |
| 57 | Specific Phobia | - | LRZ |
| 58 | GAD, MDE | - | ECP |
| 59 | MDE, Anxiety NOS, MDE | GAD, MDE, AUD, PD+AG | - |
| 60 | - | MDE | - |
| 61 | PTSD, GAD, MDE, DYS | Eating Disorder | VEN, MPD, LRZ, PZN, QTP, GBP, BPN, EZP |
| 62 | - | OCD, GAD | ECP |

Subjects 1-23 were evaluated using a SCID-I for DSM-IV-TR, while subjects 24-62 were evaluated using the SCID-I for DSM-5. Clinically-salient depression was defined as any subject with diagnoses of BPAD with MDE, DEP NOS, DYS, and/or MDE. Clinically-salient anxiety was defined as AG, ANX NOS, GAD, OCD, PD ± AG, SAD, Specific Phobia, and/or Social Phobia. ADHD, Attention-Deficit/Hyperactivity Disorder; AG, Agoraphobia; ANX, Anxiety; AUD, Alcohol Use Disorder; BPAD, Bipolar Affective Disorder; DEP, Depression; DYS, Dysthymia; ETOH-Abuse, Alcohol Abuse; GAD, Generalized Anxiety Disorder; MDE, Major Depressive Episode; NOS, not otherwise specified; OCD, Obsessive Compulsive Disorder; PD+AG, Panic Disorder with Agoraphobia; PD-AG, Panic Disorder without Agoraphobia; PTSD, Post-Traumatic Stress Disorder; SAD, Social Anxiety Disorder; APM / DXAM, Amphetamine / Dextroamphetamine; APR, Aripiprazole; APZ, Alprazolam; BPN, Buprenorphine-naloxone; BSP, Buspirone; BUP, Bupropion; CLN, Clonidine; CLP, Clonazepam; CTP, Citalopram; DLX, Duloxetine; ECP, Escitalopram; EZP, Eszopiclone; FLX, Fluoxetine; GBP, Gabapentin; LDA, Lisdexamfetamine; LRZ, Lorazepam; LTG, Lamotrigine; LTM, Lithium; MIR, Mirtazapine; MLT, Melatonin; MPD, Methylphenidate; PAX, Paroxetine; PZN, Prazosin; QTP, Quetiapine; SERT, Sertraline; TPM, Topiramate; TZD, Trazodone; VEN, Venlafaxine; ZPD, Ziprasidone.

**Supplementary Table 3. Comparing the top three healthy control gradients from the present study to those previously published in Katsumi et al. (2023).**

|  | **Gradient 1**  Present study | **Gradient 2**  Present Study | **Gradient 3**  Present Study |
| --- | --- | --- | --- |
| **Gradient 1**  Katsumi et al (2023) | **0.692** | 0.444 | 0.105 |
| **Gradient 2**  Katsumi et al (2023) | 0.414 | **0.673** | 0.653 |
| **Gradient 3**  Katsumi et al (2023) | 0.280 | 0.130 | **0.741** |

Values reflect Pearson correlation coefficients for comparisons between healthy control gradients reported in the present study and those reported in Katsumi et al. (2023), which used data from healthy young adult participants (*n*=1003) from the Human Connectome Project.

**Supplementary Table 4. Demographic and psychometric characteristics of functional motor disorder (FND-motor) and functional seizure (FND-seiz) subgroups.**

|  | **FND-motor** (N = 49)  Mean ± SD or N | **FND-seiz** (N = 24)  Mean ± SD or N |
| --- | --- | --- |
| **Age (years)** | 41.3 ± 13.3 | 37.6 ± 15.7 |
| **Sex** | F: 43; M: 6 | F: 18; M: 6 |
| **SDQ-20** | 35.5 ± 13.1 | 35.1 ± 10.7 |
| **PHQ-15** | 13.5 ± 6.7 | 13.2 ± 5.5 |
| **BDI-II** | 17.0 ± 12.1 | 16.2 ± 12.3 |
| **STAI-Total** | 82.0 ± 22.4 | 81.4 ± 25.7 |
| **PCL-5** | 28.6 ± 18.9 | 25.4 ± 18.1 |
| **CTQ-Abuse** | 31.8 ± 13.2 | 31.0 ± 13.4 |
| **CTQ-Neglect** | 21.0 ± 8.7 | 20.7 ± 9.8 |
| **SSRI/SNRI** | 24 | 13 |

F, Female, M, Male; SDQ-20, Somatoform Dissociation Questionnaire-20; PHQ-15, Patient Health Questionnaire-15; BDI-II, Beck Depression Inventory-II; STAI-Total, Spielberger State-Trait Anxiety Inventory-Total; PCL-5, Post-Traumatic Stress Disorder Checklist for DSM-5; CTQ, Childhood Trauma Questionnaire; SSRI/SNRI, selective serotonin reuptake inhibitor/serotonin norepinephrine reuptake inhibitor use.
