## Supplementary figures and images for "Functional Connectivity Gradients Reveal Altered Hierarchical Cortical Organization in Functional Neurological Disorder"

### Supplementary Figure 1

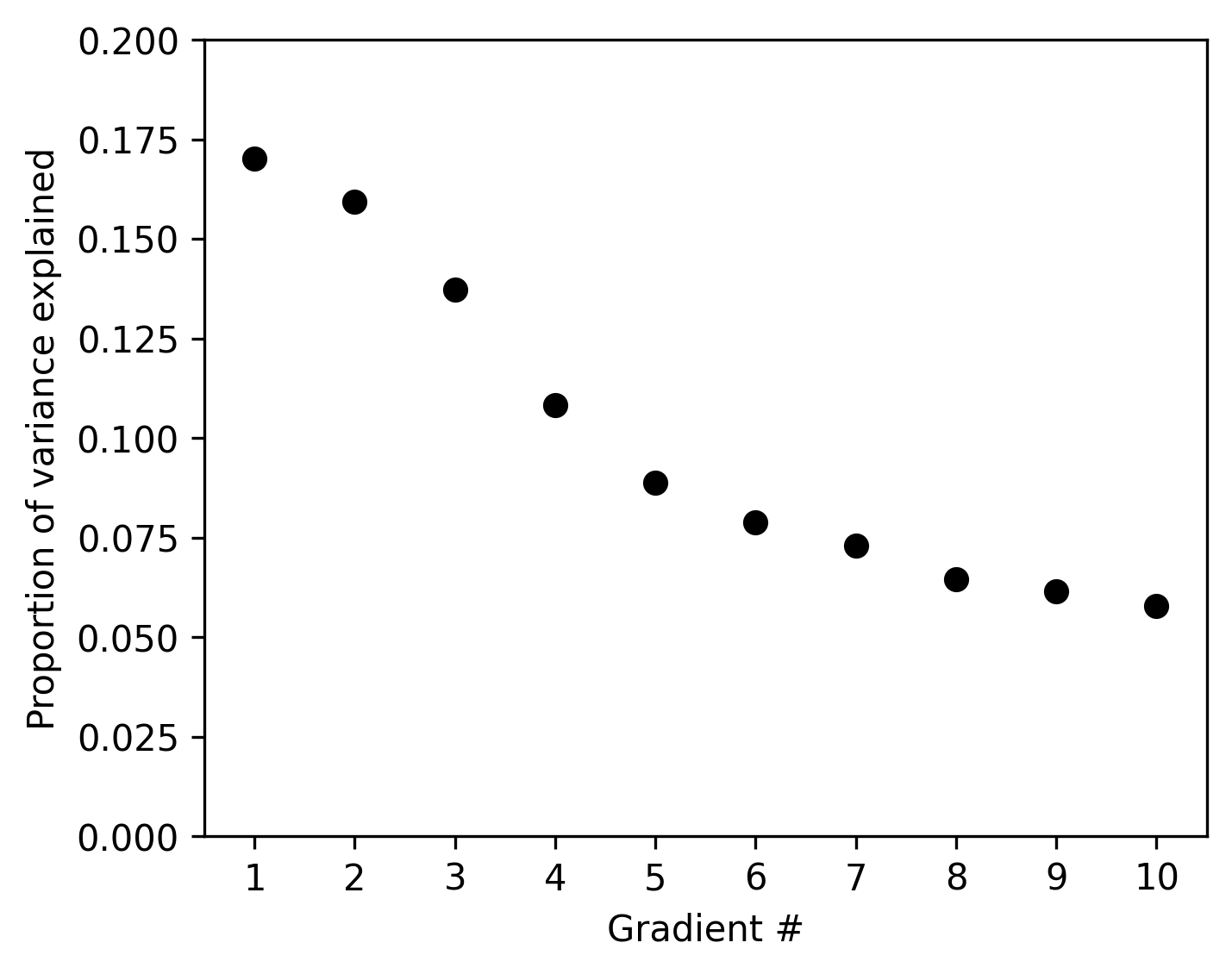

### Supplementary Figure 2

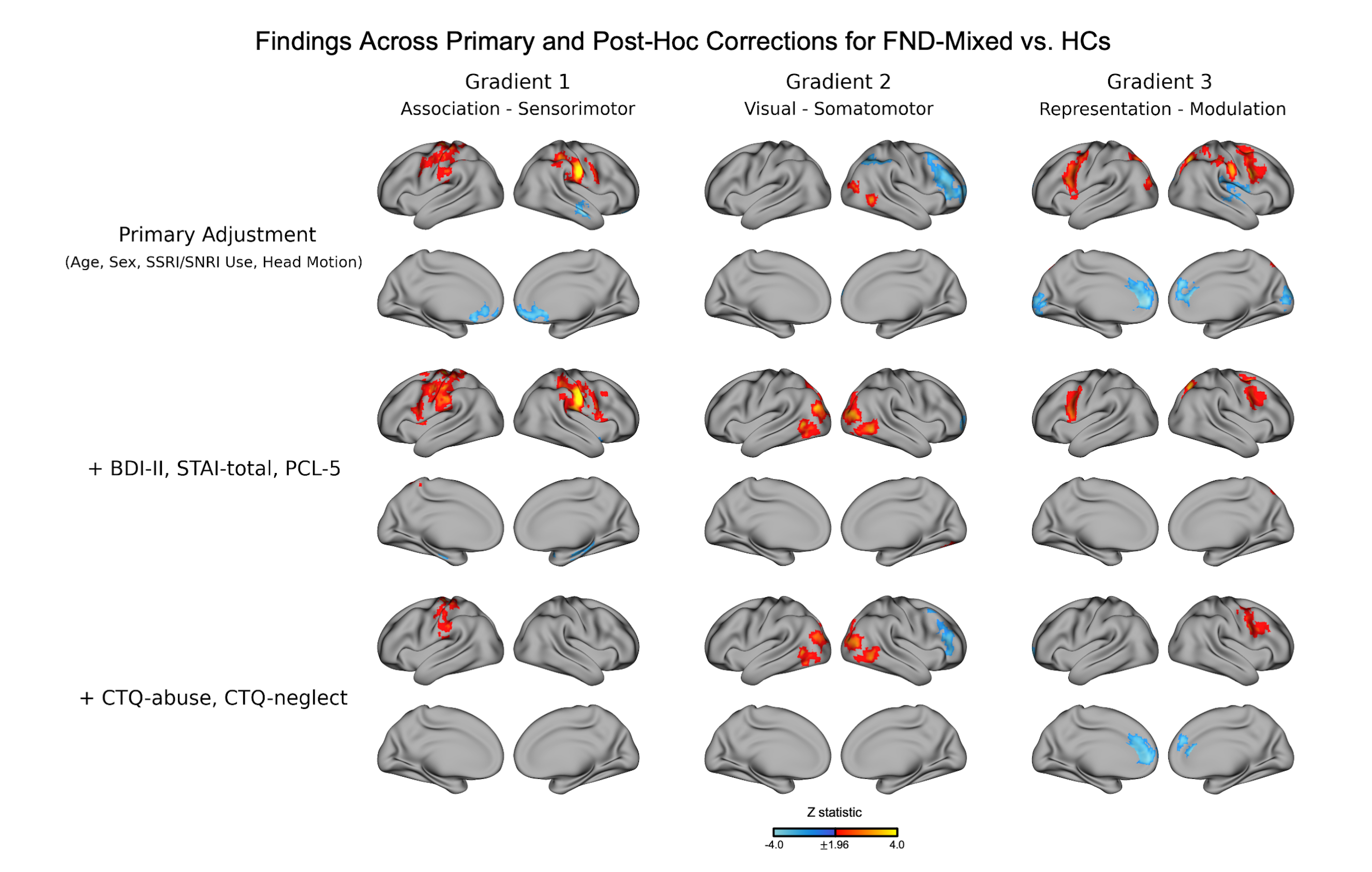

### Supplementary Figure 3

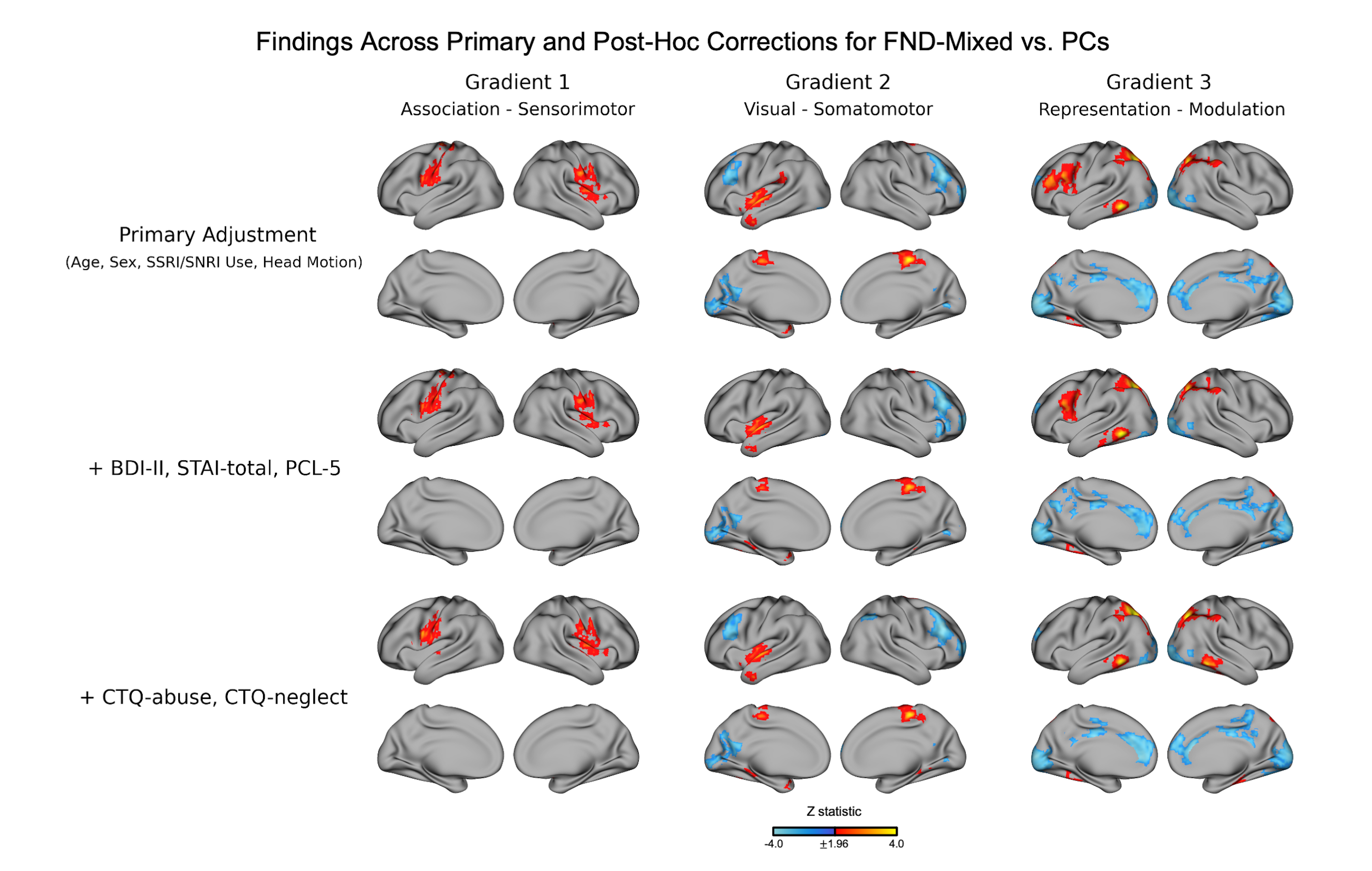
